# A wealth of opportunities: an ethnographic study on learning to deliver high-value, cost-conscious care

**DOI:** 10.1101/19011916

**Authors:** Lorette A. Stammen, Linda M.E. Janssen, Guusje Bressers, Erik W. Driessen, Laurents P.S. Stassen, Renée E. Stalmeijer, Fedde Scheele

**Affiliations:** family medicine resident and PhD-candidate in the field of Medical Education, School of Health Professions Education (SHE), Department of Educational Research and Development, Maastricht University, Maastricht Limburg, The Netherlands; recently graduated physician, currently working in geriatric health care. At time of the study, she was a graduate student at the faculty of health, medicine and life sciences, Maastricht University, Limburg, The Netherlands; anthropologist and PhD-candidate in the field of Medical education, School of Health Professions Education (SHE), Department of Educational Research and Development, Maastricht University, Maastricht, Limburg, The Netherlands; professor of Medical Education, Department of Educational Research and Development, Maastricht University, Maastricht, Limburg, The Netherlands; professor of Medical Education and a gastro-intestinal surgeon, Department of Surgery, Maastricht University Medical Center, Maastricht, The Netherlands; assistant professor, and qualitative researcher, School of Health Professions Education (SHE), Department of Educational Research and Development, Maastricht University, Maastricht, Limburg, The Netherlands; professor of Health Systems Innovation and Medical Education at the Athena Institute, VU School of Medical Sciences, Amsterdam UMC, Amsterdam, The Netherlands and a gynaecologist at the OLVG hospital Amsterdam, Amsterdam, The Netherlands

**Author notes:** **Corresponding author:** Lorette A. Stammen, MD, Department of Educational Development and Research, Faculty of Health, Medicine and Life Sciences, Maastricht University, PO Box 616, 6200 MD Maastricht, The Netherlands.

**Keywords:** medical education, high-value, cost-conscious care, residency training, ethnography

## Abstract

**Objective:** Since physicians’ behaviour determines up to 80% of total healthcare expenditures, training residents to deliver high-value, cost-conscious care is essential. Residents acknowledge the importance of high-value, cost-conscious care-delivery, yet perceive training to be insufficient. We designed an observational study to gain insight into how the workplace setting relates to residents’ high-value, cost-conscious care-delivery.

**Design:** This ethnographic study builds on 175 hours of non-participant observations including informal interviews, 9 semi-structured interviews and document analysis.

**Setting:** Department of obstetrics and gynaecology in an academic hospital in the Netherlands. Population or sample: 21 gynaecology residents.

**Methods:** Iterative analysis process of fieldnotes, interview transcripts and documents, including open-coding, thematic analysis and axial analysis by a multidisciplinary research team.

**Results:** Residents rarely consider health care costs, and knowledge regarding costs is often absent. Senior consultants guide residents while balancing benefits, risks and costs, with or without explicating their decision-making process. Identified learning opportunities are elaboration on questions raised concerning high-value, cost-conscious care, checking information about costs that are used in discussions about high-value, cost-conscious care, and having a more open and explicit discussion about high-value, cost-conscious care.

**Conclusion:** Our study emphasizes that the opportunities and potential to train residents to deliver high-value, cost-conscious care in the workplace are present. The challenge resides in capitalizing on these opportunities. We suggest departments to consult external experts to facilitate discussions regarding high-value, cost-conscious care to contribute to informal learning and to create a workplace setting in which high-value, cost-conscious care-delivery is prioritized.

**Funding:** none

## Introduction

Rising healthcare expenditure significantly impact the sustainability of healthcare systems.^1-3^ Limiting costs whilst ensuring the quality of care is therefore high on the political and medical agenda.^4^ Since physicians’ behaviour determines up to an estimated 80% of total healthcare expenditures, shaping the behaviour of future physicians is a potentially valuable intervention for ensuring delivery of high-value, cost-conscious care (HV3C).^5-9^ HV3C is defined as ‘care that aims to assess the benefits, harms, and costs of interventions’ that leads to ‘care that adds value’ for the individual patient and the population in general.^10^ HV3C is characterized by three key principles: 1) it avoids wasteful care that has no added value for the patient, therewith reducing unnecessary costs; 2) it aligns with the personal preferences and needs of the patient; and 3) it is based on the latest evidence.^2,10-12^

Campaigns such as *Choosing Wisely*^13^ and *Less is More*^14^ have advanced awareness of unnecessary treatments and procedures and have led to a variety of initiatives, including educational interventions. High-profile organizations such as the Accreditation Council for Graduate Medical Education (ACGME), the Canadian Medical Education Directives for Specialists (CanMEDS), together with American College of Physicians (ACP) and General Medical Council (GMC) also aim to prepare residents for their future roles in HV3C delivery.^15-17^ A general aim of these organizations is to incorporate HV3C in the training of physicians, residents and medical students.^3,16^ While initiatives mainly focused on formal education,^8,18^ residency training takes place in the clinical workplace where training is mostly informal.^19^

Workplace based learning in the area of HV3C is increasingly investigated^9,20^ and results indicate that residents perceive HV3C training to be insufficient.^9^ These findings are worrying, since workplace based learning covers the majority of residency training ^21^ and lessons from graduate medical education persist long into practice.^22-24^ We aimed to build on and expand this research by obtaining a real-life perspective on the workplace setting. Therefore, we designed an observational study to gain insight into how the workplace setting is related to residents’ HV3-delivery.

## Methods

### Research design

We conducted an ethnographic study to gain insight into how the workplace setting is related to residents’ HV3C-delivery. Ethnography is a qualitative research methodology, commonly used to study cultures, by which data is collected through fieldwork.^25,26^ In this case fieldwork consisted of non-participatory observations, informal interviews, semi-structured interviews and document analysis to generate insight into the sample population’s views, motives and actions in the real-life setting.^25,27^ For a more elaborate description of ethnography as a methodology and insight in the iterative process of data-collection and data-analysis, we would like to refer to Appendix A.

### Setting

The fieldwork was conducted in an academic hospital in the Netherlands at a Department of Obstetrics and Gynaecology. At the time of the study, a national government-subsidized project was running aimed at increasing awareness of HV3C during residency training ^28^. This project provided educational material and e-learning opportunities to stimulate residents to voluntarily set up projects to improve HV3C delivery in their department. During our data-collection no formal education was present to teach residents HV3C-delivery.

### Subjects

All 21 resident physicians working in the gynaecology department at the time of our study participated. Prior to the study they were informed about the research by letter and attended a short presentation during the morning handover, in which we explained the primary research goal as ‘we want to gain insight into how the workplace setting is related to residents’ HV3C-delivery’. Written informed consent was obtained from all participating residents. The Ethical Review Board of the Netherlands Association for Medical Education approved this study on August 24, 2017, under file number 881.

### Data collection

The second author (LMJ) shadowed gynaecology residents in daily practice for 175 hours during the period of three months. LMJ accompanied the residents during patient consultations, surgical procedures, deliveries and medical rounds, and attended meetings, e.g. morning reports, educational activities and (multidisciplinary) seminars, as well as lunch and coffee breaks. These non-participant observations (i.e. the researcher was not involved in care delivery) focused on residents’ behaviour, conversations and situations regarding care delivery. Observations were followed by unstructured and ad hoc informal interviews with the aim of gaining thorough comprehension of the context and rationale of the observed moments.^29,30^ Additionally, semi-structured interviews were held (by LMJ and LAS) with nine purposively selected participants to gain additional information about specific observed situations and the department in general from representatives of the observed population. Participants were sampled based on the level of involvement in discussions and conversations regarding HV3C, both residents and staff-members with high and low levels of involvement were selected. All interviews were audio-recorded and transcribed verbatim. Document analysis of hospital and national protocols and patient charts was executed to see, when residents or supervisors referred to protocols, whether their claims were supported as well as whether the decision-making process was retraceable.

Observed behaviour and conversations were thickly described, written up in detailed, context-sensitive and locally informed fieldnotes about the observed events^25,31^. Data collection involved iterative cycles of data collection and analysis, combining observations, interviews and analysis together with the research team (see Appendix B).^25,30^ The iterative process of data collection discontinued when data-saturation was reached, meaning data-analysis did not result in new concepts and an adequate understanding of key concepts was gained.^26^ Data saturation was reached after approximately 175 hours of observation and 9 semi-structured interviews.

### Data-analysis

As is common in ethnography, the data were analysed iteratively (see Appendix B) starting with line-by-line open coding by LMJ, LAS and GB.^25,32^ The next steps were thematic and axial analyses, in which we identified central themes and relationships between the various themes. As a result of team discussions, we gradually adapted the focus of the observational themes and conducted semi-structured interviews in order to deeper our understanding of the data. Additionally, FS checked if the observations represented national care delivery and not a specific regional setting. We are aware of the influence of the researchers’ background on the process of data-collection and data-analysis, to increase transparency we included a reflexivity-paragraph in Appendix A.

## Results

### The consideration of benefits, risks and health care costs in daily practice

When protocols, guidelines or regional agreements failed to provide clear direction towards care delivery, or when new scientific information became accessible, health care professionals needed to decide which care delivery was considered *‘appropriate’*. Health care professionals then considered pros and cons, including concerns for patients, consequences of a missed diagnosis, respect for autonomy of patients, burden of testing or treatment, influence of quality of life and long-term health benefits. We rarely observed attention for health care costs, or conversations regarding who would eventually pay for those costs. In cases in which costs were included in the process of balancing pros and cons of health care services, the absence of knowledge regarding costs became apparent. Questions regarding financial costs arose when residents and senior consultants discussed a scientific article, reviewed the protocol, or discussed the necessity of structural testing. Observations demonstrated that financial reasoning did not influence the process of decision making. We did not observe occasions in which health care costs were checked as a result of such discussions.

*It is the end of the morning round and medical students, supervised by the resident-in-training are critically appraising a procedure. The resident noticed differences between the value and use of repetitive PCR-ratio* (*protein-to-creatine*) *determined in single voided urine versus ACR-ratio in 24-hour-collected urine for diagnosing pre-eclampsia* (*PE*) *in their training region*.

*O13: Actually, this article shows that there is little difference between PCR* (*protein-to-creatine ratio*) *and ACR* (*24-hour albumin-to-creatinine ratio*).

*S3: And what about the cost?*

*A26: That’s a good one, we should actually check that in the lab*.

*Document analysis demonstrated that costs were not checked nor was the protocol adapted as a result of the discussion*. (*Fieldnotes, February 21, 2018*)

### The influence of the senior consultant on the process of balancing benefits, risks and costs

HV3C-delivery by residents was guided by senior consultants’ preference. In some cases, the senior consultant extensively ‘walked residents through’ the process of balancing all relevant stakes that led to their proposed diagnostic or treatment plan. In other situations, the senior consultant stated their opinion of what they considered to be appropriate care without further explanation. Although residents not always understood nor agreed with the senior consultant, in general they did not request insight in the senior consultant’ considerations regarding what was deemed appropriate. Residents tended to execute the advice of the senior consultant with little countering of the consultants’ judgment.

Variation between senior consultants was prominent and recognizable for residents and was sometimes considered confusing especially in the eyes of junior residents. Senior residents used these differences to get agreement on their own proposed plan by being deliberate in whose senior consultant’s advice they sought in certain cases, as mentioned by resident R17 in an informal interview.

> *R17: ‘There are pretty big differences between the senior consultants. As a resident you have to get to know the bosses, so you know how they work. At some point you know the preferences of every senior physician, and if you have a certain plan in mind, you just call the senior consultant that you think is likely to agree with your plan*… *Sometimes you think it is nonsense to do something and then you just need a senior consultant who agrees with you*.*’*

> (*Fieldnotes, February 22, 2018*)

### Three observed learning opportunities regarding the process of balancing benefits, risks and health care costs

Based on the analysis of our observations, informal and formal interviews, it became apparent that there were three main areas for learning which could be improved. First, although residents and staff-members raised questions regarding what constitutes high-value, cost-conscious care, we observed that questions regularly remained unanswered or undeliberated. In these situations, conversations were discontinued (for example by raising a different question) or paused (*‘we should check that’*), without revisiting raised questions or checking/changing the course of action. Our observations also demonstrated questions followed by (collective) discussions, explicating the process of balancing benefits, risks and costs of care delivery. We asked both residents and supervisors how questions related to care delivery could be enhanced to facilitate residents’ learning. Residents shared that they were in need of a more structural review of patient cases, followed by an organized overview of benefits, risks and costs of the case at hand. This could be done for example during the morning handover when discussing a patient case. Secondly, decision making often was based on assumptions about costs without efforts to check them. This was observed for example when costs were considered *‘high’* yet actual amounts were not known or the meaning of commonly used terminology (for example what do we mean by *‘bedrest’*) was not understood or defined. Thirdly, face-to-face conversations about care delivery were absent in instances where the line of reasoning was not clear for either staff or residents. Although senior consultants were sometimes openly critical to residents about how colleagues balanced benefits, risks and costs of care delivery, this did not result in a face-to-face conversation about the delivered care and consequently did not result in a take-home message for residents. For example, critiques often ended with comments such as; *“they probably had a good think about it”* or *“let’s continue with the next case”*. Supervisors confirmed that especially formal meetings created interesting teaching opportunities but were hesitant to deepen the discussion in those meetings. Supervisors motivated this by describing how they aimed to maintain a safe learning environment (do not give the resident the impression their decision was *‘false’*) while also exposing residents to the uncertainty that is present in care delivery (modelling that there is more than one *‘correct’* treatment decision).

## Discussion

### Summarising

Training residents to provide HV3C is essential for the sustainability of healthcare systems.^2,5,9,10^ This study explored how the workplace setting is related to the residents’ HV3C-delivery. Our results demonstrated that costs are not structurally considered in the process of care delivery. Another finding was the strong influence of the senior consultants’ opinion regarding what is considered high-value, cost-conscious care on the resident. Furthermore, learning opportunities regarding HV3C were identified; elaboration on questions raised concerning HV3C, checking information about costs that are used in discussions about HV3C, and having a more open and explicit discussion about HV3C.

### Insight in relevant health care costs

The importance of knowledge-transmission for the training of high-value, cost-conscious care delivery has been emphasized in previous research.^7,9^ We observed that physicians aim to make health care costs part of the discussion, yet lack information to do so highlighting the importance of knowledge of health care costs in particular. Besides the lack of formal education, another known hurdle for knowledge transmission is the complexity of health care economics and the lack of transparency of costs in health care systems.^33^

### The consultant as leading role model

The leading role of the senior consultant in health care delivery in general,^9,19,20^ and HV3C specifically^9,33^ has been previously identified. Through observing role models’ care delivery residents learn professional behaviours, particularly from those senior consultants that match their personal views.^34^ Data-analysis demonstrates that senior residents intentionally seek advice from senior consultants who are likely to share their views on HV3C-delivery. Training opportunities lie in pursuing a workplace setting where senior consultants deliberately assist residents in explicating their reasoning regarding HV3C, with or without sharing the same view. This can be achieved by inviting residents to discuss, assess and reflect on future and past care delivery. Practice variations between consultants and departments could function as a starting point for these discussions and reflections,^35,36^ especially when residents express that they doubt if care delivery meets HV3C standards.

### Workplace culture

Our results point to the absence of a culture that allows to openly question the line of reasoning of colleagues. Residents’ and staff’s tendency to favour agreement over critical discussions could explain this finding. Research findings indeed indicate that striving for harmony may interfere with the quality of post-graduate medical education and quality of care.^37^ Specifically, engaging in constructive discussions with senior consultants may be challenging in a workplace setting that is considered to be hierarchical.^38^ It is not surprising that the current workplace setting is not familiar with discussing care delivery from a HV3C perspective since issues of accessibility and affordably of health care are recent. Nevertheless, current budgetary crises in most health care systems urge for brisk culture change. To initiate a culture change, we suggest focussing interventions on senior consultants and other staff members. Changing the overarching culture and stimulating residents to actively engage in HV3C is desirable. Therefore, future research should investigate how departments can be supported in developing a critical attitude towards care delivery in light of HV3C. Action research^39^ can be a suitable strategy to challenge conventional thinking about HV3C in the clinical setting. In such design, integrating an independent external expert to regularly question care delivery, and guide group discussions in an open and tolerant manner, may speed up consciousness of value for patients, costs and the urge for bottom up system change. This ‘devils’ advocate’ could help to fill in the opportunities detected in this study; elaboration on questions raised concerning HV3C, checking information about costs that is used in discussions about HV3C, and having a more open and explicit discussion about HV3C. The ultimate achievement would be that staff members and residents would take over this role and challenge each other to underpin their decision-making process in HV3C or structurally reflect on HV3C-delivery via group-discussions.

### Strengths & limitations

The main strength of this study is the use of ethnography to gain insight in how the workplace setting is related to residents’ HV3C-delivery. A major strength of ethnography is the ability to observe things that those who are routinely involved may not see as well as collecting data that participant might not be willing to share with researchers.^26^ Another strength is the rigorous performance of our data collection and analysis, in compliance with quality criteria of qualitative research. Our study has several limitations. Participating residents might have been more favourable towards HV3C-delivery, as they were aware of the researchers’ presence and the research goal, and observations might be an overestimation of the role of HV3C in residency training. A second limitation is the restricted scope of our ethnographic research, since the observations solely took place within one single specialty in one single hospital. We aimed to mitigate this limitation by providing a description of the setting in which the research was performed. Additionally, our findings were assessed by a member of our research team who is familiar with the context but not a member of it. Nevertheless, extrapolation to other residency programs or departments must be done prudently.

## Conclusions

Our study emphasizes that the opportunities and potential to train residents to deliver high-value, cost-conscious care in the workplace are present. The challenge resides in capitalizing on these opportunities. We suggest departments to consult external experts to facilitate discussions regarding high-value, cost-conscious care to contribute to informal learning and to create a workplace setting in which high-value, cost-conscious care-delivery is prioritized.

## Data Availability

I declare that all data collected for this research is saved. There are not supplementary materials available online.

## Acknowledgements

The research team would like to thank the participating department for their cooperation.

## Disclosure of interest

nothing to declare.

## Contribution to Authorship

All assigned co-authors fulfilled their duties as described in the

## Details of Ethics Approval

The Ethical Review Board of the Netherlands Association for Medical Education (NVMO) approved this study on August 24, 2017, under file number 881.

## Funding

None

## Table/Figure Caption List

Appendix A

Appendix B

## Notes

### Competing Interest Statement

The authors have declared no competing interest.

